# Exploring best practice governance for EU-funded health research consortia: A qualitative study protocol for insight and ideation

**DOI:** 10.1101/2023.12.07.23299676

**Authors:** Ana Renker-Darby, Till Bärnighausen, Christine Neumann, Raman Preet, Marina Treskova

**Author notes:** corresponding author. (AR).

## Abstract

**Background:** International consortia have emerged as a common model to organise and fund large-scale, multi-disciplinary research in contemporary health and biomedical science. The diversity of participants, size and complexity of these consortia necessitates effective governance to achieve their research aims and societal impact. For health research consortia funded by the European Union, certain governance structures and processes have emerged out of convention. However, there is limited scientific evidence to support their use, and little is known about consortia participants’ perspectives on how governance structures could be improved to better serve the implementation of research. In this paper, we present a protocol for a qualitative study to explore the perspectives of participants in European Union-funded health research consortia on the value of governance structures and how they might be improved.

**Methods and analysis:** We will conduct a qualitative study using in-depth interviews with participants in health research consortia funded by the European Union. We will recruit participants following a purposeful sampling approach, and recruitment will continue until saturation is reached. During in-depth interviews, we will ask participants about how the governance of consortia is structured and what is working well or poorly about those governance structures from their perspective. We will draw on design thinking methods to help participants to ideate improvements in governance structures. Data will be analysed using a thematic analysis approach.

**Discussion:** Findings from this study will provide valuable evidence for developing and formulating governance structures for health research consortia. The findings of this work may also contribute to guidelines for consortium proposal submissions.

## Introduction

International consortia have emerged as a common model to organize and fund large-scale, multi-disciplinary research in contemporary health and biomedical science [1, 2]. Consortia involve collaboration between several institutions, both academic and non-academic, across multiple geographic locations and with a variety of disciplinary expertise. Such consortia are usually funded for a specific purpose over a limited funding period with a fixed-term research contract [1].

Funding agencies are increasingly promoting multi-disciplinary project teams due to increasing scientific specialization in combination with the growing need to address complex, multi-faceted societal challenges such as pandemic prevention and preparedness or climate change and health research [3]. International research consortia can provide wide-ranging expertise and interdisciplinary collaboration to understand these complex issues, as well as bringing together stakeholders across multiple geographic locations to address global problems [4].

The scale and complexity of research consortia means that they require effective governance to achieve their research aims. Governance refers to the sustained and focused attempts to direct behaviour, with the intention of producing a broadly identified outcome [3]. Governance comprises both the structures and processes through which decisions about goals, priorities and activities are made, as well as who makes those decisions. Ultimately, consortium governance should enable the project’s activities to be carried out in a way that balances the interests of the different partners and leads to the attainment of the consortium objectives [3].

Effective governance is necessary for several reasons. First, increasing scientific specialization and a growing need to address complex, multi-faceted challenges mean that consortia often incorporate researchers from a wide range of disciplines to avoid siloed approaches [3]. This diversity of backgrounds and approaches sometimes leads to challenges in the collective identification of issues and solutions [4]. Research teams are also becoming larger, more geographically dispersed, and include researchers from multiple institutions in different countries and cities [3]. Consortia also often involve increasing numbers of public-private partnerships, bringing together academic institutions with non-governmental organisations and commercial companies [5]. Each of these institutions often have different and sometimes competing goals from the research, which need to be addressed and managed during the implementation [3, 6]. Clear rules about data access, publication and patenting are also required, particularly with increasingly large datasets [3, 6]. Finally, consortia governance is required to ensure compliance with external regulations around data protection and ethics, such as the General Data Protection Regulation [6]. As such, the governance of large research consortia facilitates the implementation of the research and ensures the realisation of expected outcomes and pathways to societal impact.

Health research consortia that are funded by the European Union (EU) have varying governance structures depending on their aims, but at a high level there are certain commonalities. Governance structures are partially determined by the funding body, both in the requirements for submitting grant proposals and through the contractual agreement between the consortium and the funder. For example, EU-funded research requires that consortia are structured in work packages, each with their own milestones and deliverables. Each grant agreement confirms the timelines for deliverables as well as reporting periods and mechanisms [3]. Governance structures must also conform to EU policies, such as those of gender equality and inclusiveness [7]. Other aspects of consortia governance structures are determined by the external regulations that the consortium is bound by, including data protection laws, ethics, and regulations of the various participating institutions [1, 3].

Other key elements of governance structures have emerged through convention. Consortia typically employ modular components to coordinate the activities of partner institutions and align them with the funding agency priorities [3]. These often include a management team or steering committee responsible for the internal governance of the research consortia. Consortia usually also have varying external advisory bodies that provide high-level governance for the consortium’s direction. Consortia use different external advisory bodies depending on their aims, but this often includes an ethics advisory board and scientific committee of external experts. These explicitly defined governance structures are complemented by a range of informal governance processes that are used in practice and vary between consortia.

Participation in the governance structures of consortia often demands significant time from the consortia participants. Previous qualitative interviews with consortia participants found that consortium governance structures can create additional bureaucratic workload that can delay scientific work [5]. As such, it is crucial that both the governance structures and processes of consortia support participants to implement the consortium’s research activities and meet research objectives outlined in the grant agreement without creating unnecessary work that can tie up participants’ expertise and resources [1, 8].

While there is some previous empirical research on the governance of health research consortia, this has tended to focus on the power dynamics between institutions from high-income countries and low- or middle-income countries [9-13] or the development of data governance policies within health research consortia [6, 8, 14]. Further literature draws on authors’ experience within consortia to explore the way that conventional governance structures can undermine scientists’ ability to implement responsible research and innovation [3], or to propose principles or guidance for consortia governance [1, 4]. To our knowledge, there is currently no empirical research that explores the value of governance structures from the perspective of consortia participants across a range of health research consortia.

Further, literature on how governance structures could be improved is usually limited to abstract principles or guidelines. At present, we lack research that draws on the perspectives of consortia participants to ideate improvements in governance structures. Design thinking, a human-centred approach for innovating solutions, offers methods for ideating concrete solutions to complex problems in health research [15, 16]. Design thinking has been previously employed in healthcare research, but has rarely been used in other areas of health research [15]. Design thinking methods offer an innovative approach for ideating improvements in consortia governance from the perspective of participants.

In this study, we aim to understand the perspectives of consortia participants on how the governance of EU-funded health research consortia is structured, the value of governance structures and processes, and how governance structures and processes could be improved.

## Materials and methods

### Study design

This is a qualitative research study addressing four research questions:

1. How are the internal steering functions and external advisory functions of EU-funded health research consortia commonly structured?
2. What is the value of the internal steering functions and external advisory functions in EU-funded health research consortia from the perspective of consortia participants?
3. What is the value of ethics advisory boards for research in EU-funded health research consortia from the perspective of consortia participants?
4. According to consortia participants, how can the governance of EU-funded health research consortia be improved?

This study will take place between October 2023 and July 2024. The data collection method for this research is in-depth interviews with participants in EU-funded health research consortia. In the interviews, we will explore the perspectives of participants on the value of consortia governance structures for the implementation of research. We will also employ design thinking methods to ideate improvements in consortia governance structures.

### Population

The study population includes participants in active EU-funded health research consortia. Criteria for inclusion in the study are as follows:

- 18 years or older
- English speaking
- Participation in an EU-funded health research consortium that has been active for at least six months
- Member of at least one of four participant groups: i) lead scientists/researchers (overall lead, work package lead or co-lead); ii) non-lead scientists/researchers; iii) non-scientists (e.g. non-governmental organisation participants); iv) project co-ordinators or administrators.
- Agree to participate and sign the consent form

Participants who do not meet the inclusion criteria will be excluded.

### Sample size

A minimum of 20 participants will be interviewed, with at least five participants in each of the four participant groups described above and a balance of genders across participants. Recruitment and data collection will continue alongside data analysis until data saturation has been reached. We will use the concept of saturation as defined by Morse to determine when data collection will end [17, 18]. Morse describes saturation as when data from several participants have essential characteristics in common, enabling abstraction from individual participants’ data and theoretical development.

### Participant recruitment and sampling

For recruitment, we will use a purposeful sampling approach that aims to select information-rich cases for in-depth study [19]. Sampling will occur on the basis of maximum variation to collect a diverse range of perspectives and ensure transferability of findings across EU-funded health research consortia [19]. To ensure diversity of participants we will apply four criteria: participant role within the consortium (lead scientists/researchers; non-lead scientists/researchers; non-scientists; project co-ordinators or administrators), gender, country of institution, and consortium.

The research team will initially invite at least one potential participant from each group from their own networks to participate in the study. We will provide participants with a participant information sheet and consent form via email. Participants will have the chance to read the participant information sheet and ask the research team any questions before signing the consent form and emailing it to the research team. We will then schedule an interview time with the participant.

Further, we will ask the participants whether they can recommend other potential participants from their own networks in any of the four groups and to prioritise participants of a different gender to their own, who work at an institution in a different country, and who participate in a consortium that the primary institution of the study investigators is not involved in. This recruitment approach will continue until saturation is reached [18].

### Data collection

In-depth interviews will be conducted by one researcher (AR) using a semi-structured interview schedule to guide the conversation. We chose this method of data collection to collect rich, qualitative data on the subjective perspectives of participants based on their experiences in health research consortia [20]. The interview schedule was developed based on the research questions and was piloted with two of the study co-investigators. The data generated from the pilot interviews will not be incorporated into the findings of this study.

During the interview, participants will first be asked for some demographic information (gender, country, institution/organisation, position at institution, discipline) and for information about the consortium that they are part of (name, start and end date, number of partners, location of partners, the participant’s role in the consortium). If the participant is a member of more than one consortium, they will be asked to choose one to focus on in the interview. Participants will then be asked to describe how the internal steering functions and external advisory functions of their consortium are structured.

We will use two design thinking methods to explore the value of governance structures and ideate improvements. The ‘rose, bud, thorn’ tool will be used to help participants identify what is working about governance structures, what currently is not working, and what has potential [21]. ‘How might we’ ideation questions will be used to shift participants from insight to early-stage ideation, prompting participants to suggest ways that consortia governance structures could be improved [22].

We will conduct interviews using Zoom videoconferencing software, as participants are based in a range of locations. Interviews will be audio-recorded. Recordings will be transcribed and audio-recordings will be deleted as soon as transcription is complete. We will return transcripts to participants for any edits they may wish to make.

### Data analysis

Data will be analysed by one researcher (AR) using NVivo software. Data will be analysed using the thematic analysis approach described by Braun and Clarke [20, 23]. We chose thematic analysis due to its methodological flexibility and its inductive approach, enabling the development of analytical themes that respond to the research questions [23].

Data will be inductively coded, building a bank of codes. We will then arrange these codes into descriptive themes. Descriptive themes will be organised into analytical themes that respond to the research questions. Data analysis will be an iterative process, with codes being renamed and reorganised as analysis continues and new data are collected. We will use peer debriefing between co-investigators to ensure the rigour of data analysis.

### Ethics

This study received ethics approval from the Ethics Committee of the Medical Faculty of Heidelberg University on 13.09.2023 [reference number S-516/2023]. The study is conducted under the General Data Protection Regulation. All participants will be provided with a participant information sheet explaining the study and what their participation would involve. Participants will sign an informed consent form before participating.

### Registration

This study was registered with OSF Registries on 1.11.2023 [https://doi.org/10.17605/OSF.IO/TD6QX].

## Discussion

The purpose of this study is to explore the perspectives of consortia participants on the governance structures of EU-funded health research consortia and ideate ways that governance structures can be improved. At present, there is limited evidence that supports the conventionally used governance structures in health research consortia. Participation in these governance structures is often bureaucratic and time-consuming for consortia participants. This study will provide evidence on how the governance structures of health research consortia can be improved to better serve the objectives of the consortium and the needs of consortium participants.

Findings from this study can be used by researchers when setting up future health research consortia, to develop evidence-based governance structures for their consortia. Findings may also be used by the consortium that this study is part of to improve its own governance structures. Future research could build on the study findings to develop and test best practice guidelines for the governance of EU-funded health research consortia.

### Study status

Participant recruitment is underway and data collection is expected to end in February 2024.

## Data Availability

No datasets were generated or analysed during the current study. All relevant data from this study will be made available upon study completion.

## References

1. Kaye J, Muddyman D, Smee C, Kennedy K, Bell J, UK10K. ‘Pop-up’ governance: developing internal governance frameworks for consortia: the example of UK10K. Life Sci Soc Policy. 2015;11(10):1–17.

2. Burgio MR, Ioannidis JPA, Kaminski BM, DeRycke E, Rogers S, Khury MJ, et al. Collaborative cancer epidemiology in the 21st century: the model of cancer consortia. Cancer Epidemiol Biomarkers Prev. 2013;22(12):2148–60.

3. Morrison M, Mourby M, Gowans H, Coy S, Kaye J. Governance of research consortia: challenges of implementing Responsible Research and Innovation within Europe. Life Sci Soc Policy. 2020;16(13):1–19.

4. Koelle B, Scodanibbio L, Vincent K, Harvey B, van Aalst M, Rigg S, et al. A guide to effective collaboration and learning in consortia: building resilience to rising climate risks. London, UK: Red Cross Red Crescent Climate Centre; 2019.

5. Morrison M. “A good collaboration is based on unique contributions from each side”: assessing the dynamics of collaboration in stem cell science. Life Sci Soc Policy. 2017;13(7):1–20.

6. Teare HJA, de Masi F, Banasik K, Barnett A, Herrgard S, Jablonka B, et al. The governance structure for data access in the DIRECT consortium: an innovative medicines initiative (IMI) project. Life Sci Soc Policy. 2018;14(20):1–17.

7. European Commission. European research area policy agenda - overview of actions for the period 2022-2024. Brussels; 2021.

8. Morrison M, Klein C, Clemann N, Collier DA, Hardy J, Heißerer B, et al. StemBANCC: governing access to material and data in a large stem cell research consortium. Stem Cell Rev Rep. 2015;11:681–7.

9. Pratt B, Hyder AA. Governance of transnational global health research consortia and health equity. Am J Bioeth. 2016;16(10):29–45.

10. Tagoe N MS, Pulford S, Murunga VI, Kinyanjui S. Managing health research capacity strengthening consortia: a systematised review of the published literature. BMJ Global Health. 2019;4:1–12.

11. Tagoe N, Pulford J, Kinyanjui S, Molyneux S. A framework for managing health research capacity strengthening consortia: addressing tensions and enhancing capacity outcomes. BMJ Glob Health. 2022;7(e009472):1–28.

12. Pratt B, Hyder AA. Governance of global health research consortia: sharing sovereignty and resources within Future Health Systems. Soc Sci Med. 2017;174:113–21.

13. Parker M, Bull S. Ethics in collaborative global health research networks. Clin Ethics. 2009;4:165–8.

14. Muenzen KD, Amendola LM, Kauffman TL, Mittendorf KF, Bensen JT, Chen F, et al. Lessons learned and recommendations for data coordination in collaborative research: The CSER consortium experience. HGG Adv. 2022;3(100120):1–15.

15. Oliveira M, Zancul E, Fleury AL. Design thinking as an approach for innovation in healthcare: systematic review and research avenues. BMJ Innov. 2020;7:491–8.

16. Brown T. Design thinking. Harv Bus Rev. 2008;Jun.

17. Morse JM. Editorial: The significant of saturation. Qual Health Res. 1995;5(2):147–9.

18. Morse JM. “Data were saturated…”. Qual Health Res. 2015;25(5):587–8.

19. Patton M. Qualitative evaluation and research methods. Beverly Hills, CA: Sage; 1990.

20. Braun V, Clarke V. Successful Qualitative Research. London: Sage; 2013.

21. The Luma Institute [Internet]. A taxanomy of innovation. Harv Bus Rev [Internet]. 2014 [cited 2023 October 10]. Available from: https://hbr.org/2014/01/a-taxonomy-of-innovation.

22. IDEO.org. The field guide to human-centred design. Canada: IDEO.org; 2015.

23. Braun V, Clarke V. Using thematic analysis in psychology. Qual Res in Psychol. 2006;3(2):77–101.

